# COVID-19: A Chronological Review of the Neurological Repercussions – What do We Know by May, 2020?

**DOI:** 10.1101/2020.05.19.20107102

**Authors:** Paulo Afonso Mei, Laura Loeb

## Abstract

**Introduction:** Despite the new SARS-CoV-2 (COVID-19) be the seventh of the coronavirus family viruses known to cause human disease, little is known about potential symptoms and syndromes secondary to the compromise of the central and peripheral nervous systems. We reviewed neurological manifestations due to the new coronavirus, thus far published in the literature, as well as guidelines issued by sub-specialties in Neurology, to tackle the disease.

**Methods:** we searched medical databases, such as PubMed, PubMed Central, LILACS and Google scholar for papers (case reports, short letters, case series, etc) describing neurological symptoms in patients with confirmed or suspect COVID-19 diagnosis and also searched webpages of associations and organizations that deal with neurological disorders.

**Results:** we describe briefly each article considered for this review. Forty-one papers were found associating neurological conditions and COVID-19. Cases are divided by disease groups and, within each disease group, results are listed in chronological order or publication date. We also discuss briefly recommendations for neurological patients, according to disease group.

**Conclusion:** Although there is evidence of neurological manifestations with previous coronaviruses, COVID-19 is assuring a volume of published papers not seen before for other coronavirus infections. Most neurological cases are not life-threatening, but 10 to 20% of cases will require hospitalization and are in risk for sequelae and death. Although a lot of data coming from these papers is amassing, researchers must bear in mind that many papers currently published are not yet peer-reviewed, and thus are prone to further corrections.

## Introduction

It’s been more than a century since a worldwide panic caused by a virus spread occurred, when of the influenza outbreak in 1918. Since <u>December 2019</u>, a new coronavirus strain, that first appeared in the province of Hubei, China (1), has raised alertness to the same level as of during the Spanish flu pandemics, due to the ease of transmission, associated with a non-negligible estimated mortality rate of more than 3% (2).

The newly discovered virus, known as 2019-nCoV, COVID-19, or SARS-CoV-2 (the last two nomenclatures are preferred in this article and are equally used) - SARS an acronym for Severe Acute Respiratory Syndrome, is the seventh in a list of coronaviruses, all from the *Orthocoronavirinae* sub-family, that can cause harm to humans. Two coronavirus of the alpha genera, HCoV-NL63 and HCoV-229E, and four coronaviruses from the beta genera, HCoV-OC43, HCoV-HKU1, SARS-CoV (or SARS-CoV-1) and MERS-CoV, complete the list.

As with previous strains, SARS-CoV-2 is a positive-sense, single-stranded RNA virus that causes mainly mild respiratory symptoms that can go unnoticed, as the vast majority of cases are a- or oligosymptomatic, but in a considerable fraction of patients, it can result in more severe illness. World Health Organization (WHO) official data shows that COVID-19 has currently infected more than 4.5 million people and accounts for more than 300,000 fatalities worldwide (3), more than 14,000 only in Brazil. Diagnosis can be confirmed by either molecular assay by polymerase chain reaction, PCR, or by detection of immunoglobulin antibodies – IgM, IgA, and IgG – against protein N. PCR remains the gold standard and is performed, specifically, in the case of this RNA virus, through reverse transcriptase, or RT-PCR (for practical purposes, we opted for the term PCR in this article), usually acquired from a nasal and oropharyngeal swab, or by bronchoalveolar lavage, although it can be performed in any tissue or fluid, including cerebrospinal fluid (CSF).

There is increasing evidence that SARS-CoV-2 is a systemic infection that reaches different organs and systems, including the nervous system by either direct and/or indirect routes (4). This is largely expected, based on knowledge of previous coronavirus infections, particularly from the more recent waves of SARS (5) (6) (7) and MERS - Middle East Respiratory Syndrome (8) (9) (10) (11). Both diseases account for at least 15 case reports in the literature of neurologic complications related to infections (12). **Table 1** encompasses some examples of case reports published regarding those two viruses and their neurological complications.

Among more ancient strains, OC43 is notably associated with cases of neuronal and glial inflammation, the most severe a fatal case of encephalitis in an 11-month-old boy with congenital immunodeficiency, described in 2016 (13). Post-mortem immunohistochemical analysis allowed the identification of viral particles within the neuropil.

Central Nervous System (CNS), Peripheral Nervous System (PNS) and neurovascular conditions, like viral meningitis, anosmia/hyposmia, encephalitis end encephalopathy, Guillain-Barré Syndrome (GBS), myositis, cranial nerve palsy, stroke, and others have been linked to SARS-CoV-2, making it extremely important for health professionals to better understand the link between this pathogen and its neurological repercussions. We present a review of current data available on the neurological consequences of the new coronavirus.

## Methods

We did a systematic review of papers published in English, up to May ..14^th^._l_..202.0. PubMed and PubMed Central (14), EMBASE (15), LILACS (16), and Google Scholar (17) databases were searched. Articles meeting the association between the terms “neurology”, “nervous system”, “neurological”, “Guillain Barré”, “muscle”, “rhabdomyolysis”, “CSF”, “cerebrospinal”, “intensitve care”, “ICU”, “critical”, “EEG”, “epilepsy”, “seizures”, or “spinal fluid” AND either “COVID-19”, “SARS-CoV-2”, “coronavirus”, “2019-nCoV”, “MERS” and “SARS” were accessed. Later, we searched all those databases and included a broader search in Google (18), for “neurology”, “nervous system”, “neurological”, AND “guidelines”, “society”, or “recommendations”. For neurological cases, only papers published in scientific journals, with or without peer review, were included. For guidelines, websites of recognized scientific organizations and associations were also accepted.

## Results

We found 41 articles consisting of case reports or series of cases with nervous system compromise by the new SARS-CoV-2, plus a variety of statements and documents issued by associations, with guidelines to neurologists and patients. Articles come from a variety of countries, mainly USA, China, Iran and Italy. **Figure 1** shows all countries where articles encountered come from.

We must emphasize that, at this point, and due to the rush to gather information that current times require, the majority of articles found and cited are yet to be peer-reviewed and, therefore, modified and corrected.

We will discuss the findings below, which are separated by topics regarding disease groups affecting the CNS, PNS, and neurovascular system, as well as guidelines issued by specific organizations, to tackle COVID-19 complications. In each segment, articles are enlisted in the chronological order they’ve been published. **Table 2** displays a summary of those articles.

Readers should likewise keep in mind that we do not analyze in deep the general clinical situation of cases, as the main focus remains on neurological symptoms. However, when required, information about other systems is covered in the description.

### 1. Neurological Alterations Associated with COVID-19

In an analysis of 214 patients infected with COVID-19 in Wuhan, China (19), published in <u>February 25, 2020</u>, 78 (36.4%) manifested neurological symptoms, being dizziness (16.8%), headache (13.1%), muscular pain/injury (10.7%), impaired consciousness (7.5%), hypogeusia (5.6%) and hyposmia (5.1%) the most commonly reported.

In another series of 58 patients admitted in two Intensive Care Units (ICUs) with confirmed SARS-CoV-2 in Strasbourg, France, published on <u>April 15, 2020</u>, Helms et al. (20) verified neurologic symptoms in 49 (85%). A few patients presented these symptoms at admission, and the majority after withholding of sedation and neuromuscular blockers. Of seven patients that underwent spinal tap, only one had an elevation of Immunoglobulin G and proteins. All cases had negative results for PCR research for SARS-CoV-2 in the CSF.

We will disclose disease groups of cases so far published in the topics ahead. **Table 3** lists cases described separated by disease groups, as well as the number of patients bearing each condition.

#### 1.1 Diseases Affecting the CNS

##### Encephalitis and Encephalopathy^1^

The previously discussed analysis from the 214 patients in Wuhan (19) found one patient to manifest seizure. It is not clear, however, if it concerns one case of epilepsy of new-onset or just exacerbation of seizures in a patient that already had the diagnosis of epilepsy.

A report from Filatov et al. (21) from <u>March 21, 2020</u>, describes a Dutchman, with previous ischemic stroke secondary to Atrial Fibrillation, that was admitted in Boca Raton, FL, USA, with headache and disturbance of conscience. CSF had increased protein levels, but ordinary white and red cell counts. PCR in the spinal fluid was performed only for Herpes Simplex (HSV) and cytomegalovirus (CMV), but not for COVID-19.

Karimi and colleagues (22) report on <u>March 28, 2020</u> a female patient in Iran, aged 30, with respiratory symptoms that started 5 days prior to admission, and fever 2 days later. In the previous 48 hours before seeking attention, she had had at least five tonic-clonic seizures, and had another when being examined at the hospital, with post-ictal confusion. She did not have a history of epilepsy. The diagnosis of COVID-19 was made by nasopharyngeal PCR. Spinal tap had normal cellularity within the upper limit of normality (5 cells/μL) and normal biochemical assay. MRI of the encephalon was also unremarkable. The patient was treated with phenytoin, levetiracetam, chloroquine, and lopinavir-ritonavir, and discharged with improved health conditions one week later.

Poyiadji et al. (23), on <u>March 31, 2020</u>, depict a woman, from Detroit, MI, USA, airline worker, with headache, impairment of consciousness, and seizures. CT scan pointed to hypodensity in the medial bi-thalamic region. Subsequent MRI revealed hyperintensity in FLAIR and T2 in the same area and also in the bilateral, mesial temporal regions, while susceptibility sequences evidenced hemorrhagic spots. Hence, a diagnosis of Acute Necrotizing Hemorrhagic Encephalopathy was firmed.

On <u>April 3, 2020</u> Moriguchi et al. (24) describe a case of a 24-year-old Japanese man who was found unconscious after 9 days of initial symptoms of headache and fever that gradually worsened. He manifested seizures during emergency transportation and was admitted with a Glasgow Coma Scale of 6/15. Spinal tap evidenced pressure > 320 mmHg (elevated). CSF analysis pointed to increase WBC count (12/μL, 10 lymphomononuclear cells). PCR for SARS-CoV was performed simultaneously in the nasopharyngeal smear and in the CSF, having the latter only tested positive for the virus.

Ye and colleagues (25) describe, on <u>April 10, 2020</u>, a male subject from Wuhan, China developing confusion and alteration of conscience 13 days after having manifested dyspnea and myalgia. Brain CT was normal, and chest CT was compatible with ground glass alterations. Examination revealed meningeal irritation signs and Babinski reflex – it is not clear if uni- or bilaterally. Testing was positive for COVID-19. The spinal tap revealed an opening pressure of 220 mmHg (therefore elevated). Cerebral spinal fluid (CSF) analysis was normal – including the SARS-CoV-2 test in this fluid, which was negative. Search for bacterial and mycobacterial agents came clear.

On <u>April 15, 2020</u>, Yin et al (26) report a case of a patient treated in the large field Hospital of Huoshenshan, built by the Chinese government in a record time of 10 days (27) for the pandemics. A male, aged 64, developed lethargy alternating with irritability, 13 days after initial symptoms, which included fever and cough. PCR of the oropharyngeal swab tested positive for SARS-CoV-2. Signs of pyramidal disinhibition were present in the lower limbs. He had meningism. The patient was treated with arbidol, ribavirin, and traditional Chinese medicine. CSF obtained had normal protein level and cellularity, but opening pressure was elevated - 200 cm H_2_0, indicating elevation of the intracranial pressure. Nucleic acid test for COVID-19 in the CSF was negative. Brain CT was normal, and the authors do not disclose if an MRI was performed.

Neurologists from Brescia, Italy, report on <u>April 17, 2020</u> (28), the story of a 60-year-old man admitted with a 5-day history of worsening irritability, confusion, asthenia, and fever. Nasopharyngeal PCR confirmed COVID-19. The neurological exam was normal except for exacerbation of glabellar and palmomental reflexes and meningism. CSF displayed lymphocytosis (18 cells/μL) and hyperproteinorachia (696 mg/dL). EEG showed diffuse slowing, but no sharp waves. He was treated with Lopinavir/Ritonavir and hydroxychloroquine. CSF PCR for coronavirus and other agents came negative. Brain MRI was unremarkable. After three days and maintenance of the clinical status, he was put under intravenous methylprednisolone (1g/day for five days). A second CSF showed the same level of WBC but worsening of protein levels (1,272 mg/dL). He was discharged with the improvement of symptoms eleven days after being admitted. A third CSF showed protein levels of 914 mg/dL and 38 WBC/μL. Antibodies panel, including NMDAR, anti-Gaba receptors, CASPR2, and anti-MOG, among others, came normal.

<u>On the same date</u>, in Los Angeles, USA, Duong et al (29) depict a 41-year-old obese woman with diabetes, and complaints of headache and fever, with no respiratory symptoms. CT of the head was normal. Examination revealed photophobia and neck stiffness. CSF showed elevation of WBC – 70/μL (100% lymphocites), protein of 100 mg/dL and glucose 120 mg/dL (serum was 200). EEG showed generalized slowing. Despite normality of the CT of the chest, a test for COVID-19 was ordered (it is not specified which test), and came positive. She showed partial improvement, by the time of publication.

On <u>April 21, 2020</u>, Zhang et al. (30), from New Jersey, USA, report a female in her forties, hypertense, dyslipidemic, previously with mild respiratory symptoms and in use of azithromycin, who presented in the emergency department with dysphagia, dysarthria, headache, myalgia, and sluggishness. Examination confirmed bulbar impairment and aphasia, with left, mild facial palsy. CSF revealed normal cellularity, protein, and glucose levels. MRI showed diffuse hyperintensity in fluid-attenuated inversion recovery (FLAIR) and T2 sequences, and diffusion-weighted imaging (DWI) showed restriction in the deep white matter of the hemispheres, indicating a case of Acute Disseminated Encephalomyelitis (ADEM). Magnetic Resonance Angiography (MRA) as normal. Apparently, no MRI of the spinal cord was performed, possibly because she did not manifest pyramidal signs in the limbs. COVID-19 molecular testing was positive and was performed two days after admission.

Dugue and colleagues report, on <u>April 23, 2020</u>. (31), CNS manifestations in a 6-week-old male infant from New York, USA. He presented episodes of sustained upward gaze and bilateral leg stiffening. CSF had 2 leukocytes/μL and a protein level of 40 mg/mL. Testing for COVID-19 was positive in nasopharyngeal swab and negative in the spinal fluid.

On <u>May 1^st^, 2020</u> a paper from Sohal and Mossammat (32) describe a 72-year-old man, in New York, USA, with coronary disease, diabetes, and renal impairment, already on hemodialysis, with severe respiratory distress, requiring intubation. PCR for SARS-CoV-2 was positive. On the third day of admission he manifested multiple tonic-clonic seizures. CT of the brain showed microangiopathy, and no acute signs. EEG confirmed sharp waves in the left temporal region. On the fifth day, the patient worsened and died before CSF was collected.

Galanopoulou et al. (33) disclose, on <u>May 06 2020</u>, electrographic findings in a retrospective study in New York, USA, from EEGs performed <u>between March 1^st^, 2020 and April 15^th^</u> in 22 COVID-19 positive patients, of whom four had prior diagnosis of epilepsy. Neuroimaging had new findings in 3 of 13 of which MRI was requested (white matter hyperintensity in one, subdural hematoma in one, subarachnoid hemorrhage in one). Overall, EEGs displayed disorganized background, and 9 of 22 (40.9%) of these cases had epileptic discharges – frontal in 8/9 – bilateral and symmetric in 3/8, bilateral asymmetric in 5/8 and unilateral in 2/8.

Researchers from Lausanne, Switzerland, report on <u>May 07, 2020</u> (34), two female patients, aged 64 and 67, the first with psychosis and seizures, and the second with headache and stupor. Brain MRI was normal for both, and CSH presented lymphocytosis and was negative for PCR for SARS-CoV-2.

##### Myelitis

Zhao and other doctors of the Hubei province, China (35) depict on <u>April 09, 2020</u>, a case of a 66-year-old man with respiratory symptoms and SARS-CoV confirmed by nasopharyngeal swab PCR who later developed flaccid paraparesis with compromise of the sphincters (urinary and bowel incontinence). Neurological examination revealed sensory loss below the T10 level. Brain CT revealed chronic abnormalities (atrophy, lacunar infarction). CSF and MRI were not obtained.

#### 1.2 Diseases affecting the PNS

##### Anosmia

Anosmia rate in patients with the new coronavirus varies drastically. In the aforementioned article, Mao et al. (19) found, in Wuhan, 11 patients of 214 (5.1%), with hyposmia, and 12 (5.6%) with hypogeusia.

Bagheri et al. (36) performed an ecological study with an online questionnaire. Results were published on <u>March 27, 2020</u> More than 15,000 Iranians living within the country responded to the forms, from whom 75% reported anosmia. Although the authors only surveyed symptoms and did not test the participants, they judge that there were no other viruses massively circulating in Iran by the time of the research, and that there was a significant correlation between anosmia and COVID-19 positivity in different provinces. Their results contrasted, therefore, with reports of Mao et al of one single digit of percentage of hyposmia complainers.

Belgium authors report, on <u>April 6, 2020</u> (37), results from a survey of 417 confirmed COVID-19 cases - 263 females, 56 mild or moderate smokers, no heavy smokers (i.e., more than 20 cigarettes per day). They were required to answer a short version of the QOD-NS (Questionnaire of Olfactory Disorders-negative statements), among other questions. Anosmia or hyposmia were diagnosed in 357 (85.6%) and hypogeusia by 342 (88.8%).

In a correspondence letter by Galougahi and colleagues (38), on <u>April 11, 2020</u>, a 27- year-old male patient from Tehran, Iran, was submitted to MRI of the olfactory bulbs, after having confirmation for COVID-19 by PCR. Anosmia was the only complaint, but it is not stated how many days before imaging the symptoms had started. They state that different from this patient, it is not unusual to find reduction of olfactory bulbs volume after a viral infection, correlating to the loss of smell, and conclude that more patients need to be imaged, and if possible, with use of additional techniques, such as SPECT- MRI with Thallium-201.

A series of 114 patients in France with COVID-19, reported in <u>April 17, 2020</u>, by Klopfenstein et al (39), verified a prevalence of anosmia of 47%, which was often associated with dysgeusia.

Stratification of anosmia by degrees of smell loss was performed by Moein et al (40). In a case-control study published on <u>April 17, 2020</u>, 60 patients with COVID-19 and 60 healthy controls were asked if they felt the loss of smell, having 21 (35%) from the COVID-19 group responded yes, versus none of the controls. Then, the University of Pennsylvania Smell Identification Test (UPSIT), which tests for 40 different odors, was applied. According to UPSIT scores, anosmia was diagnosed in 15/60 patients with SARS-CoV-2 (25%), versus 0 in controls, moderate and severe microsmia, altogether, were present in 36 coronavirus patients (60%), versus 0 controls. Normosmia or mild microsmia was present in 9 COVID-19 patients (15%) and all 60 controls.

Anosmia appears to not only be an indicator of disease, but also of prognosis, as stated by Yan et al, in a paper published on <u>April 24, 2020</u> Authors from California, USA (41), report a series of 169 patients that tested positive for SARS-CoV-2, of which 128 had been given information about their olfactory and gustatory status. Adjusted analysis showed that anosmia was a strong and independent predictor of non-admission, as 68 of 102 (66.7%) non-admitted patients reported partial or total loss of smelling, versus 7 of 26 admitted patients (26.9%), whereas alterations in chest x-ray was a predictor of admission. Patients with intact smelling and gustation, and also older patients or people with diabetes were more prone to aggravation of coronavirus infection. The overall prevalence of anosmia was, therefore, 75 of 128 (58.6%).

##### Guillain Barré Syndrome (GBS)

On <u>April 1^st^, 2020</u>, Zhao et al. (42) describe a female patient, aged 61, being admitted for ascending weakness in the lower limbs and fatigue. She had recently visited the Wuhan area in China. CSF displayed normal cellularity but elevated protein level – 124 mg/dL. Blood panel also showed lymphopenia and thrombopenia.

In a letter to the editor, Toscano et al. (43) report, on <u>April 17, 2020</u>, having observed five cases of GBS in patients that were diagnosed with the new coronavirus. Neurological symptoms emerged between 5 and 10 days after the first respiratory symptoms. Three patients manifested the axonal form, two with the sensorimotor and one with the pure motor variations. The remaining two patients displayed the demyelinating form. Spinal tap in those with the axonal form revealed an increase of protein levels, between 101 and 193 mg/dL, whereas protein level was within the normal range in the demyelinating patients. White blood count in the spinal fluid was normal in all cases, which was expected. All patients received intravenous human immunoglobulin. Only one, by the time of the report, had fully recovered and had been discharged.

Italian experts report on <u>April 29, 2020</u> (44), the story of a male, aged 71, hypertense, previously treated for lung cancer, referred to the emergency department due to subacute paresthesia in the lower limbs that evolved in three days to severe, flaccid tetraparesis, without cranial nerve impairment. CT of the head was unexceptional. Spinal tap displayed mild elevation of cells - 9 leukocytes/μL, and protein – 54 mg/mL. PCR of the nasopharynx was positive, and of the CSF was negative for COVID-19. Nerve conduction showed a neuropathy of predominantly demyelinating features.

On <u>May 12, 2020</u>, Ottaviani and colleagues (45) describe a 66-year-old woman with impairment of gait and fatigue 3 days prior to admission, and a longer history (10 days) of respiratory symptoms. Nerve conduction studies (NCS) showed prolonged waves, denoting a demyelinating GBS syndrome. CSF showed no cells and protein level of 108 mg/dL, a typical protein-cytological dissociation associated with the disease. SARS-CoV was only detected in nasopharyngeal swab, but not in the spinal fluid.

##### Cranial nerve palsy and Miller Fisher Syndrome

Spanish doctors from Madrid describe on <u>April 17, 2020</u> (46), two patients with confirmed SARS-CoV-2 and neurological findings consistent with Guillain Barré variations. One male, aged 50, with signs of Miller Fisher syndrome (oculomotor palsy, ataxia, areflexia), associated with right internuclear ophthalmoparesis, and another male, aged 39, with ageusia, bilateral sixth nerve palsy, and areflexia. Authors interpreted this patient as having polyneuritis cranialis, which is thought to be a variant of GBS, although some authors consider it within the spectra of Miller Fisher subtypes. For practical and statistical purposes, we considered both cases as Miller Fisher.

Similar cases were reported on <u>May 1^st^, 2020</u>, by researchers from Weill Cornell Medical College in New York, USA (47), describing two cases: first, a 36-year-old male with left ptosis, diplopia and bilateral distal paresthesia in the lower limbs, having had 4 days before fever, cough and myalgia. Nasal swab PCR was positive for COVID-19. MRI displayed T2 hyperintensity and enlargement of the left oculomotor nerve. As he also had paresthesia and ataxia, he was diagnosed with Miller Fisher Syndrome, a variant of GBS, and was treated with immunoglobulin and hydroxychloroquine, being discharged with improvement three days later. Ganglioside panel was negative for antibodies. They also report a woman, aged 71, with painless diplopia, having had a cough and fever in the past days. MRI revealed the enhancement of both optic nerve sheaths. CSF had normal opening pressure, cellularity, and biochemistry. She was treated with hydroxychloroquine for pneumonia and later discharged with the perseverance of abduction palsy of the right eye, showing slow recovery in the following days.

##### Muscle lesion

Jin and Tong, from Wuhan, China, report on <u>March 20, 2020</u> (48), a man, aged 60, with respiratory distress and fever. The oropharyngeal swab was positive for SARS-CoV-2. Creatine kinase (CK) was normal at admission, so was kidney function, and lactate dehydrogenase (DHL) was slightly elevated – 280 U/L (upper limit 245). Nine days later, he displayed pain in the lower limbs, CK was 11,842 U/L, DHL was 2,347 U/L. He was treated with fluid therapy, lopinavir, and moxifloxacin, improving symptoms some days later.

Suwanwongse and Shabarek (49) report, on <u>April 06, 2020</u> in New York, USA, a male patient, aged 88, with lower limbs weakness and pain. He was hypertense, dyslipidemic - used statins for several years, had benign prostatic hypertrophy, mild cognitive impairment, chronic kidney disease, and congestive heart failure. CK level peaked 12,203 U/L, and later dropped gradually to normal levels.

#### 1.3 Diseases Affecting the Neurovascular System

##### Stroke

Data from 221 patients in Huazhong, China (50), described by Li et al in <u>January, 2020</u>, found a stroke to be diagnosed in 11 (5%) of these patients. Patients with stroke were older, with a mean age of 71.6 years old versus 52.1 years old of the remaining patients. Also, one case of cerebral venous thrombosis (CVT) and one case of hemorrhagic stroke were reported.

Vu et al publish, <u>on March 19, 2020</u> (51), three cases of COVID-19 that did not display respiratory symptoms, but instead had other symptoms – one a hemorrhagic stroke in the left basal ganglia, on a 30-year-old male. Brain CT performed cuts up to the apex of the lungs and could identify ground-glass abnormalities, and a molecular diagnosis of the new coronavirus was established.

Authors from the Netherlands (52) publish, on <u>April 10, 2020</u>, a cohort of 184 ICU patients with confirmed SARS-CoV-2, of which 31 had thrombotic complications – of those 3 were ischemic strokes. There are no further details on these cases specifically.

Researchers from North Italy published, on <u>April 19, 2020</u> a series of 6 patients (53) with a confirmed diagnosis for COVID-19 by nasopharyngeal PCR, on April 20, 2020, being 5 male and 1 female, age range 57-82. Four patients had ischemic stroke and two, hemorrhagic stroke. Among the ischemic cases, two were multi, bilateral infarcts, two compromised the right basal ganglia. One hemorrhage was in both hemispheres of the cerebellum, with compression of the fourth ventricle, another was in the frontal region, extending to the ventricle. By the time of publication, four had died and two were in poor condition.

Chinese doctors (54) describe 3 cases of stroke, on <u>April 23, 2020</u>. They were 2 males, one female, aged 65-70 with confirmed SARS-CoV-2 by PCR. In all, imaging was compatible with multiple, bilateral infarctions. All displayed positivity for anticardiolipin IgA antibodies and anti-β_2_-glycoprotein 1 IgA and IgG antibodies.

Over a two-week period from <u>March 23 to April .7, 2020</u>, Oxley and colleagues (55), all working at Mount Sinai Health System, in New York, NY, USA, noted an increase of admissions of stroke in young patients, having admitted 5 cases of males, ages 33 to 49, with large vessel strokes - three in the middle cerebral artery, one in the posterior cerebral artery and one in the internal carotid territories. In the previous year, the same service admitted a lower ratio of these cases, average of 0.73 patient younger than 50 years old per 2 weeks, hence in the observed time window there was almost a 7-fold increase in frequency. The article was published on <u>April 28, 2020</u>.

Avula et al (56) describe, on <u>April 28, 2020</u>, four patients in NY with confirmed COVID- 19 and stroke, 3 females (ages 80, 83, 88), one male, 73 years old. All were ischemic strokes of the middle cerebral arteries (two on the left, two on the right).

On <u>April 30, 2020</u>, Spanish specialists from Bilbao (57) describe an ischemic infarction in the left MCA territory in a 36-year-old healthcare female worker, smoker, with no other previous health conditions. CT angiography showed occlusion of the left ICA, MCA and ACA, with a free-floating coagulum of the Ascending Aorta. PCR for SARS-CoV was positive, and was performed because thoracic CT showed bilateral pneumonia and bilateral pulmonary embolism. It is not known if she complained of previous respiratory symptoms, as she was brought by ambulance, after being found unconscious in her apartment, alone. She died three days later, according to the report.

<u>On the same date</u>, Saiegh et al (58) describe two patients, both with confirmed COVID-19, the first a 31-year-old male with subarachnoid hemorrhage Hunt and Hess grade 3, and a 62-year-old woman with ischemic stroke with subsequent hemorrhagic transformation. In both cases, CSF PCR did not evidence viral replication of SARS-CoV-2.

In the article previously mentioned, Galanopoulou (33) described, on <u>May 06, 2020</u> epileptical activity on EEG of two patients, one with a subdural hematoma and one with subarachnoid hemorrhage secondary to aneurism, in patients positive for COVID-19. Other details were already explained above.

Doctors from New York, USA, describe three cases on <u>May 06, 2020</u> (59), with arterial thromboembolic complications despite prophylaxis, of which two consisted of ischemic strokes.

### 2. Guidelines for Neurologists and Patients with Neurological Conditions

In its fourth version, from <u>March 25, 2020</u> (60), the Association of British Neurologists’ guideline comprise of a great amount of orientations for neurologists, during the COVID- 19 pandemic. Guidelines are divided by Disease Groups. For patients with Multiple Sclerosis (MS), it is advised against stopping oral or injectable immunotherapies, except maybe for patients with severe SARS-CoV-2 manifestations. For patients with muscle diseases and cardiomyopathy, interruption of ACE inhibitors or other medications for cardiologic symptoms is not recommended. Also, steroids should not be suspended in Duchenne patients, and in other cases, when possible, dosages of prednisone above 10 mg per day should be revised but, in general, steroids and immunosuppressants should not be routinely discontinued. The same advice is given for patients with nerve disease, neuromuscular junction disorders and other autoimmune diseases other than MS. Degenerative diseases, like Amyotrophic Lateral Sclerosis (ALS), Stroke and Cognitive Disorders are not considered, per se, as risk factors, and patients should be considered moderate or high risk depending on their basal health – for instance, ALS patients with weakened respiratory muscles and FVC<60% should be tagged as high risk, whereas ALS with initial or mild symptoms should not.

The European Academy of Neurology has provided Scientific Panel Statements for Diseases Groups (61), as well. For epilepsy, it is stated that, this far, there is no proof that people with epilepsy are more prone to infection, nor that COVID-19 can trigger seizures. For Neurocritical care, a concern in the rise in subacute neurological complications, such as GBS and encephalitis is manifested.

As an increase in stroke incidence is a major concern, the American Heart Association has issued Temporary Emergency Guidance (62) to US Stroke Centers. Attention is drawn to the fact that Stroke patients may have asymptomatic or faint respiratory complaints, so physicians should be aware of this matter and consider testing for COVID-19 in these cases. Due to the shortage of ICU beds worldwide, it is advised that stable, stroke patients could be transferred to non-intensive care wards after completion of 24 hours of observation, if an intensive care unit bed is then required.

Also, an International Panel, from specialists in 18 countries (63), including the USA, China, Japan, France, and Brazil, among others, published on <u>May 3^rd^, 2020</u> a document, divided into 18 sections. According to the authors, an alarming 5% (confidence interval 2.8 – 8.7%) of the population infected with COVID-19 is expected to have a stroke. It also encouraged the use of neurological assessment by telemedicine, where a neurologist can’t be in locus, as TeleStroke units (64) have demonstrated equivalence to bedside consultation. The use of rt-PA (recombinant tissue Plasminogen Activator) for thrombolysis should be maintained, but extra concern should be raised, as septic patients from COVID-19 can lead to hyper- or hypocoagulability, resulting either in higher chances of unsuccessful reperfusion as well as massive intracerebral hemorrhage, as there is a risk of multiple organ failure in SARS-CoV-2,

Depression, anxiety, and sleep disorders are also major concerns. A Chinese survey (65) led by Zheo et al, with more than 7,000 people in China, found a prevalence of anxiety, depression, and sleep disruption to be, respectively, 35.1%, 20.1%, and 18.2%. Health workers were found to have the highest prevalence of sleep issues, when compared with other occupational groups. Younger age and more time spent with reading/viewing/listening to COVID-19 information was also was a risk factor for anxiety, when compared to the rest.

Specifically concerning sleep disorders, the European Academy of Cognitive Behavioral Treatment of insomnia (CBT-I) has issued a report (66) concerning recommendations during the pandemic, which include trying to keep a regular schedule, sparing some time for reflection and writing or talking about one’s feelings, and avoiding using the bedrooms for activities other than sleep and intercourse, exercising regularly, among other directions.

## Discussion

Our review could detect 41 articles regarding neurological complications and COVID-19, published from late February to mid-May, 2020 and include report of 630 cases with confirmed COVID-19 (cases that were suspected, but not confirmed, are not accounted), of those 23 (3,6%) cases of CNS harm, 564 (89,6%) cases of PNS disease and 43 (6,8%) neurovascular cases. 549 cases (87%) are of anosmia/hyposmia, which evidences that, fortunately, the vast majority of neurological complications are not life-threatening or requiring hospitalization. Notwithstanding, and considering the numbers, one or two patients of every 10 patients with neurological symptoms in SARS-CoV-2 show potentially life-threatening conditions.

Concerning chronological order, one case (2,4%) was published in February, 2020, 7 (17%) were in March, 2020, 27 (66%) in April and 6 (14,6%) till mid-May.

Most articles come from countries already struck with a considerable number of cases and deaths – mainly USA, China and West Europe. We also cite 6 articles of neurological complications of SARS-CoV-1 (or simply SARS-CoV) and MERS, and one of OC43, as examples that nervous damage by coronaviruses has been previously verified.

Despite all papers found, it remains unclear at this point if CNS and PNS damage occur directly due to neuronal invasion and destruction of the neurons and glial cells by COVID-19 or secondarily in consequence of systemic events such as cytokine storm (67). This uncertainty is reassured, when of the 22 cases reporting encephalitis or encephalopathy, and of all literature available so far, to our knowledge, only one patient tested positive, by RT-PCR, for SARS-CoV-2 in the CSF, reported by Moriguchi. All other cases had no RNA replication detectable in the spinal fluid. That includes GBS cases and in other situations where CSF was obtained (in some patients it wasn’t collected).

Of all neurologic complications, stroke is a major concern, as there is at this point a trend in the rise of neurovascular cases, especially stroke from large arteries and to a great extent in young patients. Although the pathophysiology is still not fully explained, it is known that patients with COVID-19 are prone to a hypercoagulability or dyscrasia states, in the same fashion as happens with other viruses, such as HIV (68), cytomegalovirus, varicella-zoster (69), Hepatitis C, Epstein-Barr (70), SARS-CoV (6) and MERS-CoV (10).

Of the 43 mentioned cases of stroke, 38 (88%) were ischemic, and the remaining 5 (22%) were hemorrhagic. This series is proportional to data previously described of vascular events unrelated to infections, that is, the ratio of ischemic to hemorrhagic strokes, in neurovascular services, is usually between 80 to 90% ischemic to 10-20% hemorrhagic strokes, which could raise a suspicion that, in a few cases, infection of COVID-19 could be coincidental. However, this line of though seems to be invalid for most cases, due to the undoubtful increase in cases of young people, or people without comorbidities that raise the risk of stroke, to the occurrence of multi infarctions in both hemispheres, without proven cardioembolic etiology, usually the culprit in bilateral manifestations, and to the confirmation by some author, like Kiok, of a greater than usual incidence of thrombotic complications of any sort (not only neurological).

Special attention to immediate care and, to thrombolysis or thrombectomy should be sought in cases of ischemic stroke, especially the ones from large cerebral arteries, prone to graver outcomes and death. On the other hand, the greater risk of bleedings, due to the coronavirus septic, dyscrasic state, is to be considered. Finally, higher risk of renal impairment by the multi-organ failure due to SARS-CoV-2 is worrying, especially in patients receiving iodine contrast for CTs and angiographies, and plays against optimal outcomes from stroke.

Specifically, about anosmia, it is probable that trans-synaptic (or transcribrial) invasion, proposed by Baig et al. (71), i.e., via axonal, retrograde path of the olfactory nerve, explains only partially this symptom, as many patients that display loss of smelling also manifest loss of taste (dysgeusia), because the latter has nothing to do with olfactory bulb invasion by the virus, and also to the fact that, in one case, Galougahi performed an MRI of a patient with COVID-19 and found no alterations in the olfactory bulb – more cases with imaging are needed to warrant the confirmation of no acute injury detected in the olfactory bulb. Dysgeusia could be due to edema and/or inflammation, as happens with other airborne infections, but also to direct damage of gustatory papillae (taste buds), in the same fashion of invasiveness to the olfactory nerve, by binding to ACE-2 receptors, as theorized by Finsterer and Stollberger (72).

Moreover, it is not clear how patients with anosmia could be less prone to severe forms of COVID-19, as the study by Yan et al. found. One theoretical explanation is that, in patients where trans-synaptic invasion occurs, nervous system damage by other pathways, i.e., via bloodstream or cytokine storm, as proposed by Wu et al. (73), is mitigated. However, this needs further exploration and research. Also, the prevalence of anosmia varied widely in studies, – 5.1%, 35%, 47%, 58.6% and 75%, in 5 reports – inferring that it may be possible that milder degrees of smell impairment go unnoticed, as the study leaded by Moein concludes, after confronting self-declaration and diagnosis after application of the UPSIT test, which is more reliable than self-awareness of smell loss.

Regarding GBS, it is becoming increasingly clear that neurological deficits from polyneuropathy may arise earlier than expected, to the extreme of starting before respiratory symptoms, as reported by Zhao. This case, specifically, is an odd manifestation, as the vast majority of neurological symptoms in GBS start weeks after the original infection (usually, but not exclusively, respiratory or from the gastrointestinal tract, or systemic like in Dengue and Zika viruses). This may be because nerve damage results from direct damage (para-infectious), and not from crossed immunity against components of the myelin sheath or the axon, the usual mechanism of polyneuropathy in GBS.

Nonetheless, both axonal and demyelinating forms in GBS were observed. In the case of Zhao et al.’s report, although authors consider it to be a demyelinating form, due to absence of some F waves, there is also reduction of amplitude in the antidromic sensitive conduction study of the left median and ulnar nerves, and in the motor conduction study of the left peroneal nerve, which is commonly found in axonal forms, however study was performed on day 5 of symptoms, a bit early to define more precisely, by nerve conduction techniques, the GBS form, and could be repeated later, for better clarification. Finally, Miller-Fisher syndrome, consisting of ataxia, areflexia and ophthalmoplegia (sometimes not all of those) was observed by Gutiérrez-Ortiz and Dinkin.

It is noticeable that, although there is a considerable number of guidelines and protocols proposed by sub-specialty associations, a substantial level on uncertainty is perceptible. For instance, there are doubts about maintenance of steroids and at which dosage, and the same applies to ACE-2 inhibitors, widely used by many neurological patients, such as people with muscular dystrophy, stroke and neuroimmune diseases.

## Conclusion

In this paper we proposed a chronological review of information available of 41 papers of cases and also declarations issued by neurological sub-specialties about the impact of COVID-19, regarding clinical aspects – the prevalence of neurologic symptoms, manifestations in the CNS, PNS, and neurovascular system and guidelines concerning specific handling of disease groups’ populations in Neurology.

Most of the data, so far, comes from reports from countries where COVID cases were or are significantly high – 80% of reports come from China, Italy, Iran and the USA, although reports from other countries start to emerge.

The prevalence of neurologic symptoms varies widely, so far from 36.4 to 85%, and it seems that patients with severe disease tend to display more frequent alterations of the nervous system than patients with mild or moderate cases.

Patients with a particular complaint – anosmia, seemed less prone to aggravation and need of hospitalization, as one study found that patients that were hospitalized were more frequently with intact olfactory and gustatory senses. Prevalence of this symptom varied widely – 5.1t %o 75%, in five different reports.

Data published so far points to less severity of neurological complications in 80-90% of times patients manifest a non-life-threatening neurological complaint, and the remaining might require hospitalization and are in risk of sequelae and even death. Among the latter, stroke is a major concern, as there is at this point, although still not clearly explained in terms of pathophysiology, a trend in the rise of neurovascular cases, and up to 5% of patients with COVID could manifest stroke, especially brain ischemia from large arteries, and also cases in young patients with little or no comorbidities.

With time, we expect to have a much broader learning of explanations for the causality of neurological symptoms by the new coronavirus, due to more publications and also to better strength and quality of further published papers, with more detailed and precise descriptions, which tend to be peer-reviewed and vetted. Also, recommendations from specialized organizations and associations tend to come with greater level of certainty.

Finally, acknowledging the fact that, among all patients with neurological complications, RT-PCR for COVID-19 was positive in the CSF in only one patient arises an intriguing question and needs further elaboration.

## Data Availability

All data is available in the manuscript

## Appendix 1 - Tables

**Table 1.**
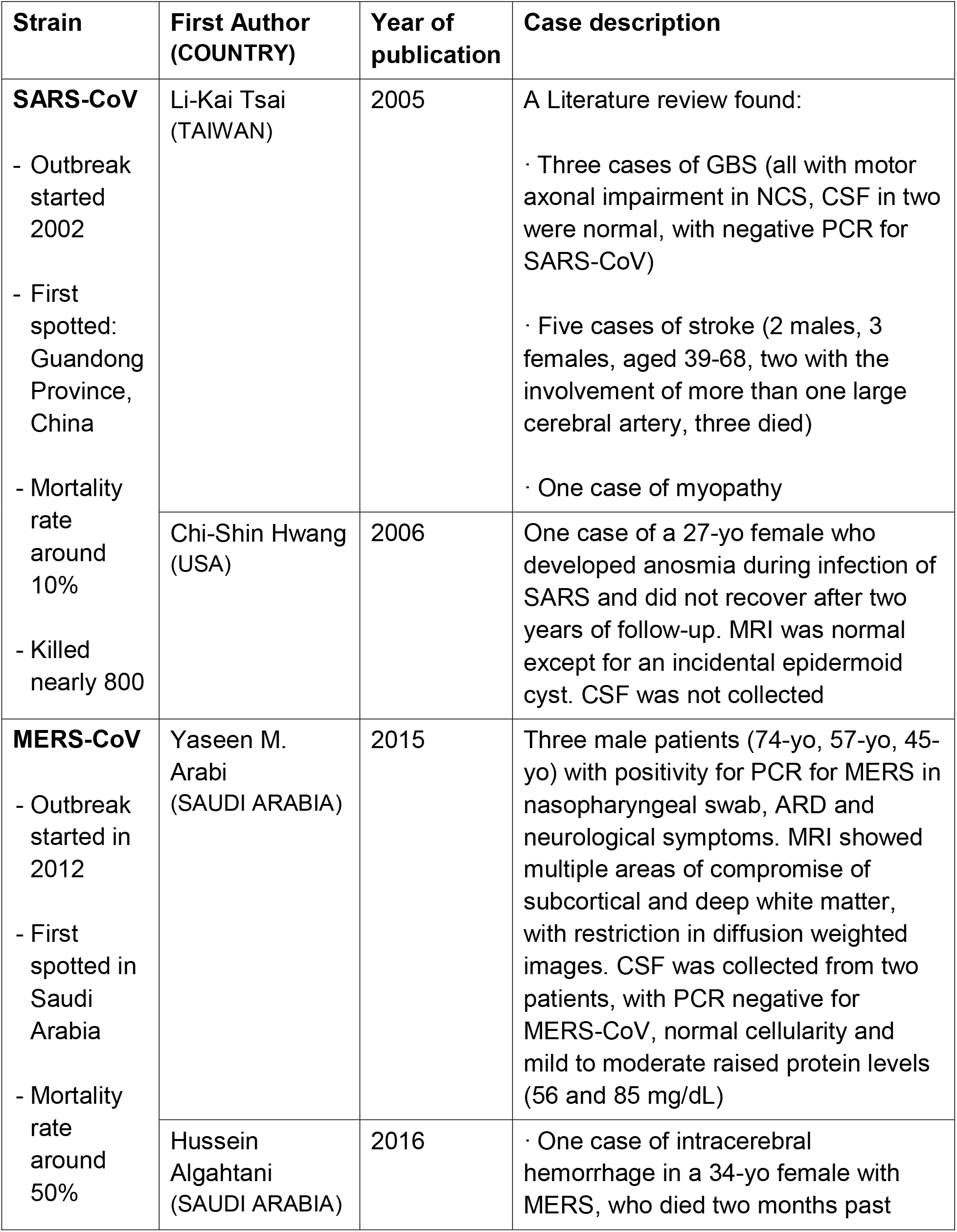
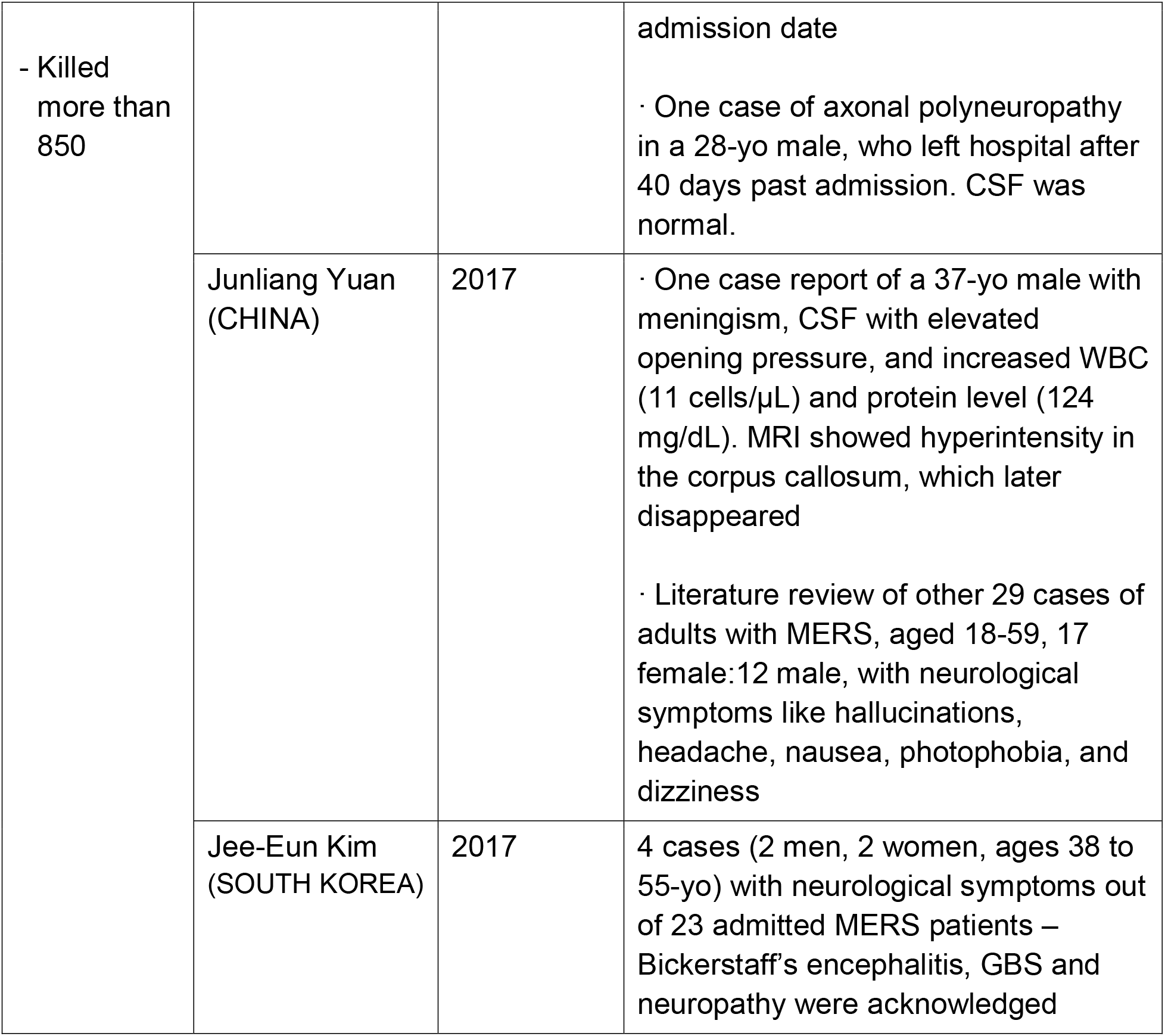
- examples of reports of compromise of the neurological system in SARS - Severe Acute Respiraroty Syndrome - and MERS - Middle East Respiratory Syndrome (yo - years-old, ARD - acute respiratory distress, CSF - cerebrospinal fluid, GBS - Guillain-Barré Syndrome, MRI - magnetic resonance imaging, NCS - nerve conduction studies, PCR - polymerase chain reaction, WBC - white blood cells)

**Table 2.** - **Timeline of Neurological Complications by COVID-19 described this far and account of papers per country** (yo = year-old, CSF - cerebrospinal fluid, CT - computed tomography, CVT - cerebral vein thrombosis, ICA - internal carotid artery, IVIg - intravenous immunoglobulin, MCA - middle cerebral artery, MRI - magnetic resonance imaging, PCA - posterior cerebral artery, PCR - polymerase chain reaction, WBC - white blood cells)

| # | Date (dd/mm/yyyy) | First author (COUNTRY) | Briefing (all cases were confirmed for COVID-19, except when otherwise stated) |
| --- | --- | --- | --- |
| 1 | 25/02/2020 | Ling Mao (CHINA) | 36.4% of 214 patients with confirmed SARS-CoV-2 in Wuhan with neurological symptoms |
| 2 | 20/03/2020 | Min Jin (CHINA) | Rhabdomyolysis with CK over 11,000 U/L in a 60-yo male |
| 3 | 21/03/2020 | Asia Filatov (USA) | Encephalitis in a 74-yo male with previous stroke of the left temporal region. EEG showed epileptic activity. CSF cellularity was normal and protein was elevated (68 mg/dL) |
| 4 | 27/03/2020 | Yanan Li (CHINA) | 11 cases of stroke (one CVT) among 221 patients (no details given) |
| 5 | 27/03/2020 | Bagheri (IRAN) | 75% prevalence of hyposmia in patients with flu-like symptoms in March (<u>OBS: patients were not tested for COVID-19, but no other viruses were reported to be in massive circulation at the time</u>) |
| 6 | 28/03/2020 | Narges Karimi (IRAN) | Female, 30-yo, with 5 tonic-clonic seizures. Brain MRI and spinal tap were normal |
| 7 | 30/03/2020 | David Vu (USA) | Hemorrhagic stroke in a 30-yo male with no respiratory symptoms |
| 8 | 31/03/2020 | Poyiadji (USA) | One case of Acute Hemorrhagic Necrotizing Encephalopathy described in a female patient. Hemorrhagic lesions are seen in the mesial bi-thalamic and bi-temporal regions |
| 9 | 01/04/2020 | Hua Zhao (CHINA) | Possible GBS associated with COVID-19 in a female patient, 61-yo; neurological symptoms, however, unusually started prior to respiratory ones |
| 10 | 01/04/2020 | M. K. Galougahi (IRAN) | MRI of olfactory bulbs from a patient with COVID-19 did not find alterations |
| 11 | 03/04/2020 | Takeshi Moriguchi (JAPAN) | 24 yo man found unconscious and with seizures, CSF showed intracranial hypertension, elevation of WBC (12 cells/μL, 10 lymphomonocytes) and positivity for SARS-CoV-2 by PCR. This is the only case, in this series, where COVID was found to replicate in the spinal fluid |
| 12 | 06/04/2020 | Kulachanya Suwanwongse (USA) | Rhabdomyolysis (CK up to ~ 12,000) in an 88-yo male with multiple comorbidities |
| 13 | 06/04/2020 | Jerome L. Lechien (BELGIUM) | 417 mild and moderate cases of COVID, anosmia and dysgeusia were present in 85.6% and 88%, respectively |
| 14 | 07/04/2020 | Thomas J. Oxley (USA) | Five patients below 50-yo with large artery strokes, three from the MCA, one from the PCA and one ICA |
| 15 | 09/04/2020 | Kang Zhao (CHINA) | 66-yo male with confirmed COVID-19 by nasopharyngeal swab, with flaccid paraparesis, T10 sensory level, and sphincter disturbance. Brain CT unremarkable, imaging of the spinal cord and CSF were not performed |
| 16 | 10/04/2020 | Mingxiang Ye (CHINA) | Wuhan male (age?) with encephalitis, normal CT and elevated opening pressure of the CSF, that had normal cellularity and biochemistry panel |
| 17 | 10/04/2020 | F. A. Kiok (NETHERLANDS) | 3 out of 184 ICU patients developed ischemic stroke; overall, 31% showed thrombotic complications |
| 18 | 11/04/2020 | Galougahi (IRAN) | MRI of a patient with SARS-CoV-2 did not show alterations in the olfactory bulbs, which were normal in intensity and volume |
| 19 | 15/04/2020 | Julie Helms (FRANCE) | 85% of patients admitted to ICUs had neurological findings |
| 20 | 15/04/2020 | R. Yin (CHINA) | 64-yo male with agitation alternating with coma, pyramidal signs present, positive for COVID-19 |
| 21 | 17/04/2020 | Gianpaolo Toscano (ITALY) | Five cases of GBS are described, being two demyelinating and three of axonal form |
| 22 | 17/04/2020 | Andrea Pilotto (ITALY) | Encephalitis with elevated WBC and protein levels in an otherwise healthy 60-yo male, EEG did not show epileptic activity. Improvement was observed after administration of IV steroids |
| 23 | 17/04/2020 | Timothée Klopfenstein (FRANCE) | Of 114 patients studied, 47% manifested anosmia, often it was associated with complaints of loss of taste |
| 24 | 17/04/2020 | Consuelo Gutiérrez-Ortiz (SPAIN) | Two cases of Miller-Fisher syndrome in two male patients, 39 and 50-yo |
| 25 | 17/04/2020 | Shima T. Moein (IRAN) | 21 of 60 patients self-reported anosmia. However, the UPSIT test detected that 51 had anosmia or moderate/severe microsmia |
| 26 | 17/04/2020 | Lisa Duong (USA) | Meningoencephalitis |
| 27 | 19/04/2020 | Mauro Morassi (USA) | Four patients with ischemic stroke, two patients with hemorrhagic stroke |
| 28 | 21/04/2020 | Tianshu Zang (USA) | ADEM in a female in her forties, MRI of the head with diffuse hyperintensity on T2 and FLAIR, CSF was normal. COVID-19 test was positive but not tested in the CSF |
| 29 | 23/04/2020 | Rachelle Dugue (USA) | 6-week-old male with upward gaze and bilateral leg stiffening. CSF was unremarkable |
| 30 | 23/04/2020 | Yan Zhang (CHINA) | Three cases of multiple, bilateral ischemic infarctions in patients with COVID-19 and positivity for anticardiolipin antibodies |
| 31 | 24/04/2020 | Carol H. Yan (USA) | 66,7% of non-admitted patients had anosmia, versus 26,9% of admitted patients |
| 32 | 28/04/2020 | Akshay Avula (USA) | Ischemic stroke in four elderly (ages 73 – 88), all from the MCA |
| 33 | 29/04/2020 | Paola Alberti (ITALY) | 71-yo male with rapidly progressing tetraparesis, CSF with mild elevation of leukocytes and protein. PCR in the CSF was negative for COVID-19 |
| 34 | 30/04/2020 | Tirso González-Pinto (SPAIN) | Massive large arteries stroke and pulmonary embolism in a 36-year-old female smoker, with confirmed SARS-CoV-2 |
| 35 | 30/04/2020 | Fadi Al Saiegh (USA) | 31-yo man with SAH and a 62-yo woman with stroke with hemorrhagic conversion, both with CSL negative for SARS-CoV-2 |
| 36 | 01/05/2020 | Sandeep Sohal (USA) | Seizures in a 72-yo man with chronic coronary and renal disease, EEG with spikes on the left temporal region, the patient passed away 5 days after admission |
| 37 | 01/05/2020 | Marc Dinkin (USA) | Ophthalmoparesis in two patients, one male, also with ataxia (Miller Fisher syndrome), one female |
| 38 | 06/05/2020 | Aristea S. Galanopoulou (USA) | Epileptic discharges were observed in 9 out of 22 patients with SARS-CoV-2 |
| 39 | 06/05/2020 | Daniel O. Griffin (USA) | Two ischemic strokes in patients that were receiving enoxaparin 40 mg subcutaneously. One in a 50-yo man (right parietal lobe), the other in a 69-yo man (left thalamus) |
| 40 | 07/05/2020 | Raphaël Bernard-Valnet (SWITZERLAND) | Two female patients with encephalopathy (seizure, psychosis, stupor). CSF with elevated WBC, PCR positive in nasopharyngeal swab and negative in CSF for COVID-19 |
| 41 | 12/05/2020 | Donatella Ottaviani (ITALY) | Demyelinating GBS in a 66-yo female, symptoms worsened despite of IVIg treatment. PCR for COVID in the CSF was negative. |
|  | PAPERS<br>PER<br>COUNTRY | USA | 16 |
|  |  | CHINA | 8 |
|  |  | IRAN | 5 |
|  |  | ITALY | 4 |
|  |  | FRANCE | 2 |
|  |  | SPAIN | 2 |
|  |  | BELGIUM | 1 |
|  |  | JAPAN | 1 |
|  |  | NETHERLANDS | 1 |
|  |  | SWITZERLAND | 1 |
|  |  | **TOTAL** | **41** |

**Table 3.**
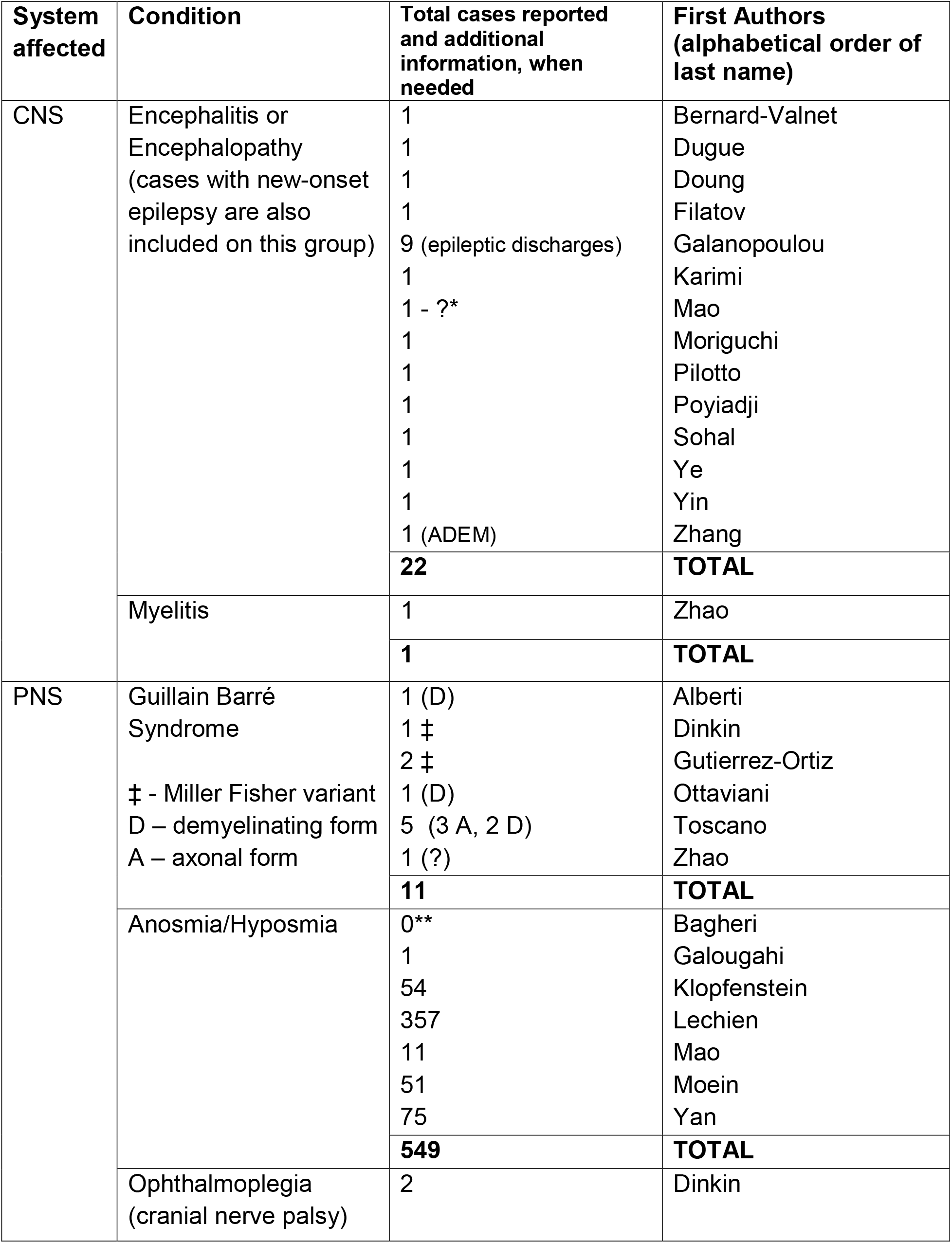
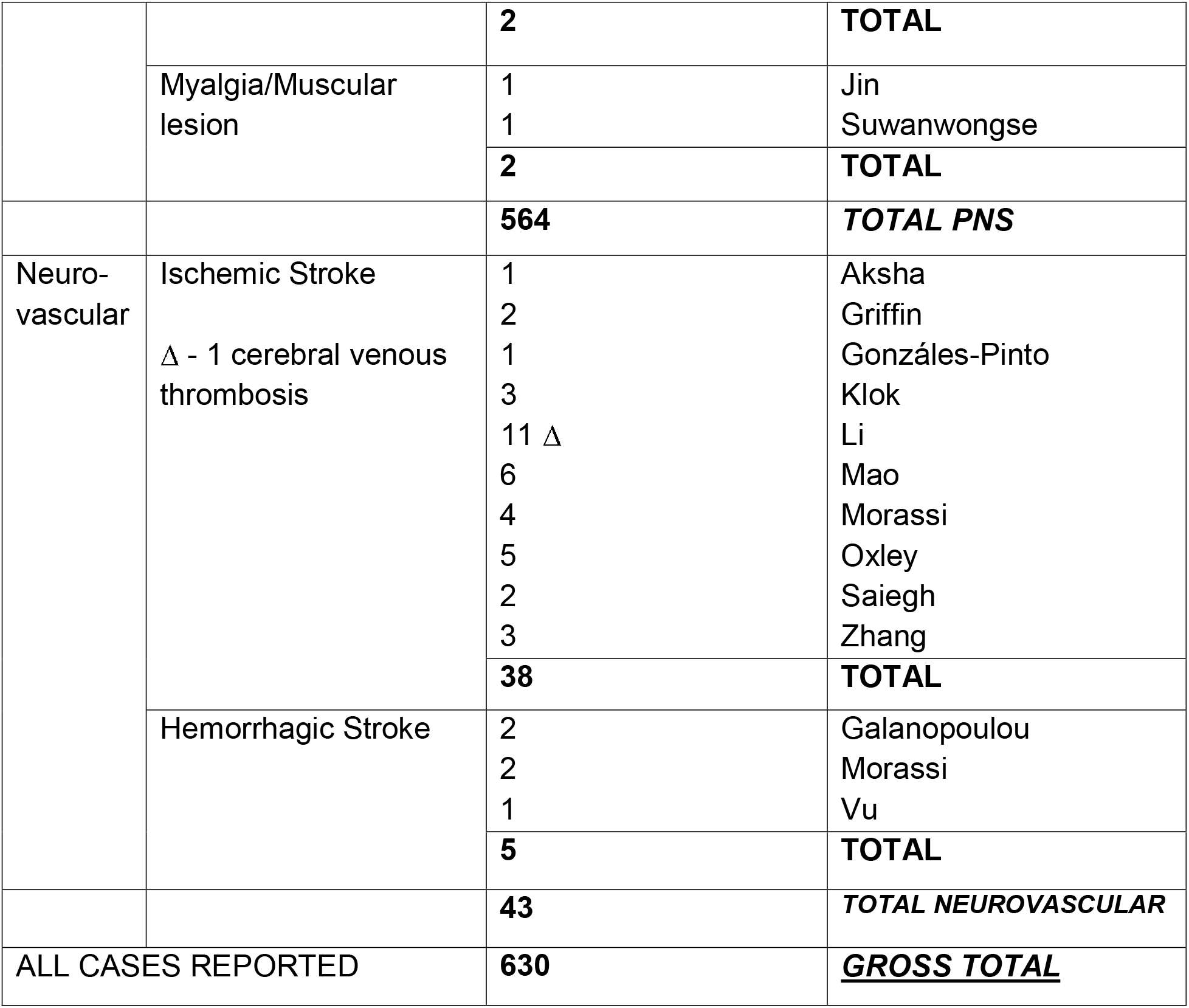
- **neurological case reported so far; divided by disease groups** (* - article describes seizure, but does not explain clearly if it meant new-onset seizure or just exacerbation of previous epilepsy; ** - we did not include the author’s results, as patients were not tested for COVID-19)

| System affected | Condition | Total cases reported and additional information, when needed | First Authors (alphabetical order of last name) |
| --- | --- | --- | --- |
| CNS | Encephalitis or Encephalopathy (cases with new-onset epilepsy are also included on this group) | 1 | Bernard-Valnet |
|  |  | 1 | Dugue |
|  |  | 1 | Doung |
|  |  | 1 | Filatov |
|  |  | 9 (epileptic discharges) | Galanopoulou |
|  |  | 1 | Karimi |
|  |  | 1 - ?* | Mao |
|  |  | 1 | Moriguchi |
|  |  | 1 | Pilotto |
|  |  | 1 | Poyiadji |
| CNS | Encephalitis or Encephalopathy (cases with new-onset epilepsy are also included on this group) | 1 | Sohal |
|  |  | 1 | Ye |
|  |  | 1 | Yin |
|  |  | 1 (ADEM) | Zhang |
|  |  | **22** | **TOTAL** |
|  | Myelitis | 1 | Zhao |
|  |  | **1** | **TOTAL** |
| PNS | Guillain Barré Syndrome<br>‡ - Miller Fisher variant<br>D – demyelinating form<br>A – axonal form | 1 (D) | Alberti |
|  |  | 1 ‡ | Dinkin |
|  |  | 2 ‡ | Gutierrez-Ortiz |
|  |  | 1 (D) | Ottaviani |
|  |  | 5 (3 A, 2 D) | Toscano |
|  |  | 1 (?) | Zhao |
|  |  | **11** | **TOTAL** |
|  | Anosmia/Hyposmia | 0** | Bagheri |
|  |  | 1 | Galougahi |
|  |  | 54 | Klopfenstein |
|  |  | 357 | Lechien |
|  |  | 11 | Mao |
|  |  | 51 | Moein |
|  |  | 75 | Yan |
|  |  | **549** | **TOTAL** |
|  | Ophthalmoplegia (cranial nerve palsy) | 2 | Dinkin |
|  |  | **2** | **TOTAL** |
|  | Myalgia/Muscular lesion | 1<br>1<br>**2** | Jin<br>Suwanwongse<br>**TOTAL** |
|  |  | **564** | **TOTAL PNS** |
| Neuro-vascular | Ischemic Stroke<br>Δ - 1 cerebral venous thrombosis | 1 | Aksha |
|  |  | 2 | Griffin |
|  |  | 1 | González-Pinto |
|  |  | 3 | Klok |
|  |  | 11 Δ | Li |
|  |  | 6 | Mao |
|  |  | 4 | Morassi |
|  |  | 5 | Oxley |
|  |  | 2 | Saiegh |
|  |  | 3 | Zhang |
|  |  | **38** | **TOTAL** |
|  | Hemorrhagic Stroke | 2 | Galanopoulou |
|  |  | 2 | Morassi |
|  |  | 1 | Vu |
|  |  | **5** | **TOTAL** |
|  |  | **43** | **TOTAL NEUROVASCULAR** |
| ALL CASES REPORTED |  | **630** | **<u>GROSS TOTAL</u>** |

## Appendix 2 - Figures

**Figure 1.**
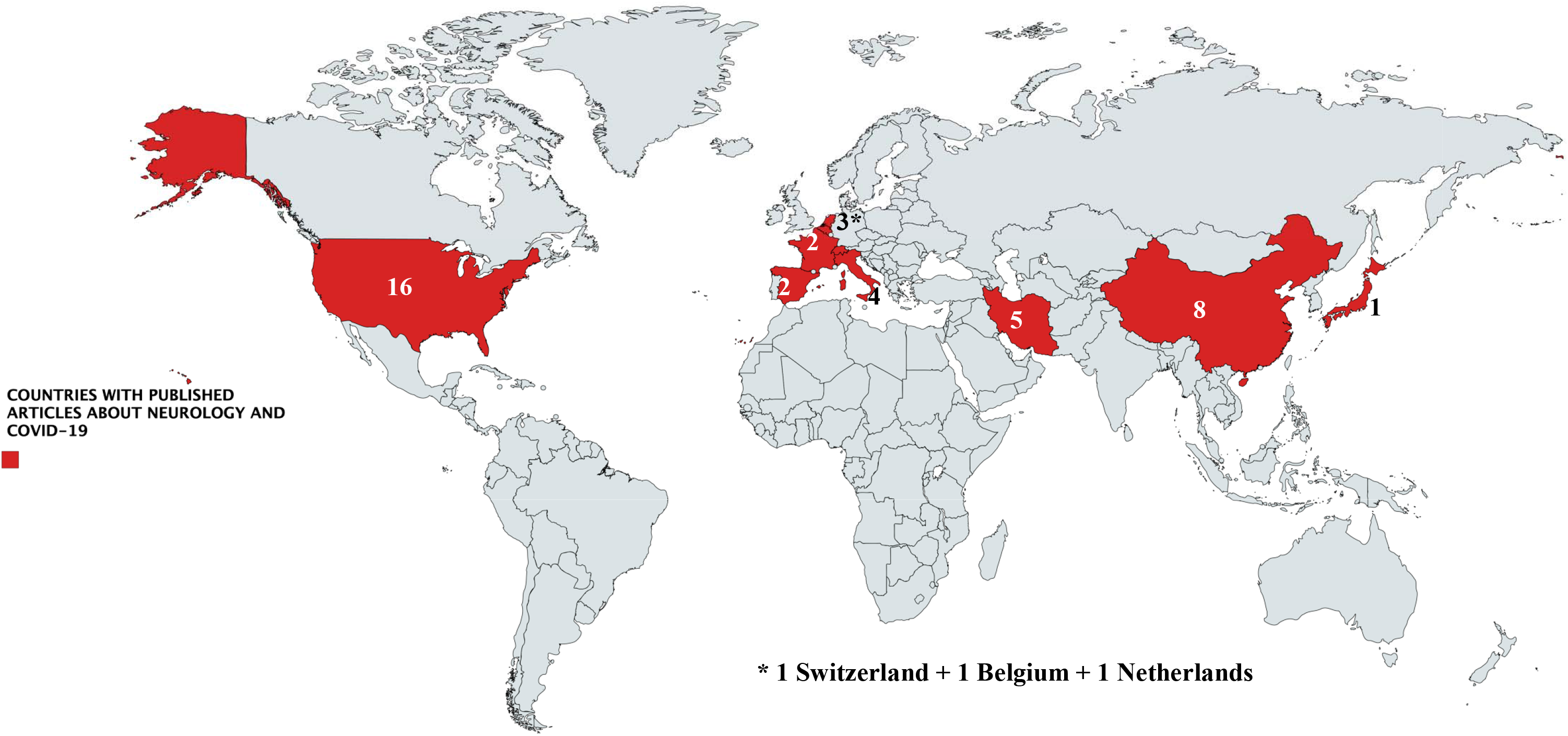
- COUNTRIES WITH NEUROLOGICAL CASES REPORTED AND NUMBER OF CASES BY COUNTRY [made with Mapchart (74)]

## Footnotes

1 - we opted to include Encephalitis and Encephalopathy on the same group, as the distinction among both entities is not always clear. Also, cases with newly-onset epilepsy were considered here, as we assume that, in patients with no past neurological issues, seizures are a direct result of viral infection, unless proven otherwise

## References

1. Zhu N, Zhang D, Wang W, Li X, Yang B, Song J, et al. A Novel Coronavirus from Patients with Pneumonia in China, 2019. N Engl J Med. 2020 Feb 20;382 (8):727–33.

2. Coronavirus Mortality Rate (COVID-19) - Worldometer [Internet]. [cited 2020 May 10]. Available from: https://www.worldometers.info/coronavirus/coronavirus-death-rate/fwho-03-03-20

3. World Health Organization. World Health Organization Covid Dashboard [Internet]. [cited 2020 Apr 25]. Available from: https://covid19.who.int

4. How COVID-19 Affects the Brain [Internet]. Medscape. [cited 2020 May 10]. Available from: http://www.medscape.com/viewarticle/928903

5. Newly discovered coronavirus as the primary cause of severe acute respiratory syndrome. - PubMed - NCBI [Internet]. [cited 2020 May 10]. Available from: https://www.ncbi.nlm.nih.gov/pubmed/12892955

6. Tsai L-K, Hsieh S-T, Chang Y-C. Neurological manifestations in severe acute respiratory syndrome. Acta Neurol Taiwanica. 2005 Sep; 14 (3):113–9.

7. Hwang C. Olfactory Neuropathy in Severe Acute Respiratory Syndrome: Report of A Case. Acta Neurol Taiwanica. 2006; 15(1):26.

8. Kim J-E, Heo J-H, Kim H, Song S, Park S-S, Park T-H, et al. Neurological Complications during Treatment of Middle East Respiratory Syndrome. J Clin Neurol Seoul Korea. 2017 Jul;13(3):227–33.

9. Yuan J, Yang S, Wang S, Qin W, Yang L, Hu W. Mild encephalitis/encephalopathy with reversible splenial lesion (MERS) in adults-a case report and literature review. BMC Neurol [Internet]. 2017 May 25 [cited 2020 May 10];17. Available from: https://www.ncbi.nlm.nih.gov/pmc/articles/PMC5445341/

10. Algahtani H, Subahi A, Shirah B. Neurological Complications of Middle East Respiratory Syndrome Coronavirus: A Report of Two Cases and Review of the Literature. Case Rep Neurol Med [Internet]. 2016 [cited 2020 May 10];2016. Available from: https://www.ncbi.nlm.nih.gov/pmc/articles/PMC4864560/

11. Arabi YM, Harthi A, Hussein J, Bouchama A, Johani S, Hajeer AH, et al. Severe neurologic syndrome associated with Middle East respiratory syndrome corona virus (MERS-CoV). Infection. 2015 Aug;43(4):495–501.

12. Ellul MA, Laura Benjamin, Bhagteshwar Singh, Suzannah Lant, MBChB, Benedict Daniel Michael, PhD, Rachel Kneen, FRCPCH, et al. Neurological Associations of COVID-19. The Lancet Neurology. Manuscript Draft. 2020 Apr 23;

13. Morfopoulou S, Brown JR, Davies EG, Anderson G, Virasami A, Qasim W, et al. Human Coronavirus OC43 Associated with Fatal Encephalitis. N Engl J Med. 2016 Aug 4;375(5):497–8.

14. PubMed [Internet]. PubMed. [cited 2020 May 6]. Available from: https://pubmed.ncbi.nlm.nih.gov/

15. Embase - Login [Internet]. [cited 2020 May 6]. Available from: https://www.embase.com/login

16. | LILACS [Internet]. [cited 2020 May 6]. Available from: https://lilacs.bvsalud.org/

17. Google Acadêmico [Internet]. [cited 2020 May 6]. Available from: https://scholar.google.com.br/

18. Google [Internet]. [cited 2020 May 13]. Available from: https://www.google.com/?client=safari

19. Mao L, Jin H, Wang M, Hu Y, Chen S, He Q, et al. Neurologic Manifestations of Hospitalized Patients With Coronavirus Disease 2019 in Wuhan, China. JAMA Neurol [Internet]. 2020 Apr 10 [cited 2020 Apr 25]; Available from: https://jamanetwork.com/journals/jamaneurology/fullarticle/2764549

20. Helms J, Kremer S, Merdji H, Clere-Jehl R, Schenck M, Kummerlen C, et al. Neurologic Features in Severe SARS-CoV-2 Infection. N Engl J Med. 2020 Apr 15;0(0):null.

21. Filatov A, Sharma P, Hindi F, Espinosa PS. Neurological Complications of Coronavirus Disease (COVID-19): Encephalopathy. Cureus [Internet]. 2020 Mar 21 [cited 2020 May 2]; Available from: https://www.cureus.com/articles/29414-neurological-complications-of-coronavirus-disease-covid-19-encephalopathy

22. Karimi N, Sharifi Razavi A, Rouhani N. Frequent Convulsive Seizures in an Adult Patient with COVID-19: A Case Report [Internet]. Vol. 22, Iranian Red Crescent Medical Journal. Kowsar; 2020 [cited 2020 May 9]. Available from: http://ircmj.com/en/articles/102828.html

23. Poyiadji N, Shahin G, Noujaim D, Stone M, Patel S, Griffith B. COVID-19-associated Acute Hemorrhagic Necrotizing Encephalopathy: CT and MRI Features. Radiology. 2020 Mar 31;201187.

24. Moriguchi T, Harii N, Goto J, Harada D, Sugawara H, Takamino J, et al. A first case of meningitis/encephalitis associated with SARS-Coronavirus-2. Int J Infect Dis. 2020 May;94:55–8.

25. Ye M, Ren Y, Lv T. Encephalitis as a clinical manifestation of COVID-19. Brain Behav Immun [Internet]. 2020 Apr 10 [cited 2020 May 4]; Available from: https://www.ncbi.nlm.nih.gov/pmc/articles/PMC7146652/

26. Yin R, Feng W, Wang T, Chen G, Wu T, Chen D, et al. Concomitant neurological symptoms observed in a patient diagnosed with coronavirus disease 2019. J Med Virol [Internet]. [cited 2020 May 5];n/a(n/a). Available from: https://onlinelibrary.wiley.com/doi/abs/10.1002/jmv.25888

27. China: Wuhan Coronavirus hospital built in just 10 days - watch [Internet]. [cited 2020 May 7]. Available from: https://www.constructionglobal.com/infrastructure/china-wuhan-coronavirus-hospital-built-just-10-days-watch

28. Steroid-responsive severe encephalopathy in SARS-CoV-2 infection | medRxiv [Internet]. [cited 2020 May 5]. Available from: https://www.medrxiv.org/content/10.1101/2020.04.12.20062646v1.article-info

29. Duong L, Xu P, Liu A. Meningoencephalitis without respiratory failure in a young female patient with COVID-19 infection in Downtown Los Angeles, early April 2020. Brain Behav Immun [Internet]. 2020 Apr 17 [cited 2020 May 12]; Available from: http://www.sciencedirect.com/science/article/pii/S0889159120305092

30. Zhang T, Rodricks MB, Hirsh E. COVID-19-Associated Acute Disseminated Encephalomyelitis: A Case Report. medRxiv. 2020 Jan 1; 2020.04.16.20068148.

31. Dugue R, Cay-Martínez KC, Thakur K, Garcia JA, Chauhan LV, Williams SH, et al. Neurologic manifestations in an infant with COVID-19. Neurology. 2020 Apr 23;

32. Sohal S, Mossammat M. COVID-19 Presenting with Seizures. IDCases. 2020 May 1;e00782.

33. Galanopoulou AS, Ferastraoaru V, Correa DJ, Cherian K, Duberstein S, Gursky J, et al. EEG findings in acutely ill patients investigated for SARS-CoV2/COVID-19: a small case series preliminary report. Epilepsia Open [Internet], [cited 2020 May 12];n/a(n/a). Available from: https://onlinelibrary.wiley.com/doi/abs/10.1002/epi4.12399

34. Bernard □ Valnet R, Pizzarotti B, Anichini A, Demars Y, Russo E, Schmidhauser M, et al. Two patients with acute meningo-encephalitis concomitant to SARS-CoV-2 infection. Eur J Neurol [Internet]. [cited 2020 May 11];n/a(n/a). Available from: https://onlinelibrary.wiley.com/doi/abs/10.1111/ene.14298

35. Zhao K, Huang J, Dai D, Feng Y, Liu L, Nie S. Acute myelitis after SARS-CoV-2 infection: a case report. medRxiv. 2020 Apr 9;2020.03.16.20035105.

36. Bagheri SHR, Asghari AM, Farhadi M, Shamshiri AR, Kabir A, Kamrava SK, et al. Coincidence of COVID-19 epidemic and olfactory dysfunction outbreak [Internet]. Otolaryngology; 2020 Mar [cited 2020 May 5]. Available from: http://medrxiv.org/lookup/doi/10.1101/2020.03.23.20041889

37. Olfactory and gustatory dysfunctions as a clinical presentation of mild-to-moderate forms of the coronavirus disease (COVID-19): a multicenter European study | SpringerLink [Internet]. [cited 2020 May 13]. Available from: https://link.springer.com/article/10.1007/s00405-020-05965-1

38. Olfactory Bulb Magnetic Resonance Imaging in SARS-CoV-2-Induced Anosmia: The First Report [Internet]. [cited 2020 May 5]. Available from: https://www.ncbi.nlm.nih.gov/pmc/articles/PMC7151240/

39. Klopfenstein T, Kadiane-Oussou NJ, Toko L, Royer P-Y, Lepiller Q, Gendrin V, et al. Features of anosmia in COVID-19. Médecine Mal Infect [Internet]. 2020 Apr 17 [cited 2020 May 6]; Available from: http://www.sciencedirect.com/science/article/pii/S0399077X20301104

40. Moein ST, Hashemian SMR, Mansourafshar B, Khorram-Tousi A, Tabarsi P, Doty RL. Smell dysfunction: a biomarker for COVID-19. Int Forum Allergy Rhinol. 2020 Apr 17;

41. Yan CH, Faraji F, Prajapati DP, Ostrander BT, DeConde AS. Self-reported olfactory loss associates with outpatient clinical course in Covid-19. Int Forum Allergy Rhinol. 2020 Apr 24;

42. Zhao H, Shen D, Zhou H, Liu J, Chen S. Guillain-Barré syndrome associated with SARS-CoV-2 infection: causality or coincidence? Lancet Neurol. 2020 May;19(5):383–4.

43. Toscano G, Palmerini F, Ravaglia S, Ruiz L, Invernizzi P, Cuzzoni MG, et al. Guillain–Barré Syndrome Associated with SARS-CoV-2. N Engl J Med. 2020 Apr 17;NEJMc2009191.

44. Alberti P, Beretta S, Piatti M, Karantzoulis A, Piatti ML, Santoro P, et al. Guillain-Barré syndrome related to COVID-19 infection. Neurol - Neuroimmunol Neuroinflammation. 2020 Jul;7(4):e741.

45. Ottaviani D, Boso F, Tranquillini E, Gapeni I, Pedrotti G, Cozzio S, et al. Early Guillain-Barré syndrome in coronavirus disease 2019 (COVID-19): a case report from an Italian COVID-hospital. Neurol Sci [Internet]. 2020 May 12 [cited 2020 May 13]; Available from: https://doi.org/10.1007/s10072-020-04449-8

46. Miller Fisher Syndrome and polyneuritis cranialis in COVID-19 | Neurology [Internet]. [cited 2020 May 11]. Available from: https://n.neurology.org/content/early/2020/04/17/WNL.0000000000009619

47. Dinkin M, Gao V, Kahan J, Bobker S, Simonetto M, Wechsler P, et al. COVID-19 presenting with ophthalmoparesis from cranial nerve palsy. Neurology. 2020 May 1;10.1212/WNL.0000000000009700.

48. Jin M, Tong Q. Early Release - Rhabdomyolysis as Potential Late Complication Associated with COVID-19 - Volume 26, Number 7—July 2020 - Emerging Infectious Diseases journal - CDC. [cited 2020 May 11]; Available from: https://www.nc.cdc.gov/eid/article/26/7/20-0445_article

49. Suwanwongse K, Shabarek N, K S, N S. Rhabdomyolysis as a Presentation of 2019 Novel Coronavirus Disease. Cureus J Med Sci [Internet]. 2020 Apr 6 [cited 2020 May 12];12(4). Available from: https://www.cureus.com/articles/30228-rhabdomyolysis-as-a-presentation-of-2019-novel-coronavirus-disease

50. Li Y, Wang M, Zhou Y, Chang J, Xian Y, Mao L, et al. Acute Cerebrovascular Disease Following COVID-19: A Single Center, Retrospective, Observational Study [Internet]. Rochester, NY: Social Science Research Network; 2020 Mar [cited 2020 May 4]. Report No.: ID 3550025. Available from: https://papers.ssrn.com/abstract=3550025

51. Vu D, Ruggiero M, Choi WS, Masri D, Flyer M, Shyknevsky I, et al. Three unsuspected CT diagnoses of COVID-19. Emerg Radiol. 2020;1–4.

52. Klok FA, Kruip MJHA, van der Meer NJM, Arbous MS, Gommers D a. MPJ, Kant KM, et al. Incidence of thrombotic complications in critically ill ICU patients with COVID-19. Thromb Res. 2020 Apr 10;

53. M M, D B, M C, S D, Gl G, C B, et al. Cerebrovascular complications in patients with SARS-CoV-2 infection: Case series. 2020 Apr 20 [cited 2020 May 11]; Available from: https://europepmc.org/article/ppr/ppr152878

54. Coagulopathy and Antiphospholipid Antibodies in Patients with Covid-19 | NEJM [Internet]. [cited 2020 May 13]. Available from: https://www.nejm.org/doi/full/10.1056/NEJMc2007575

55. Oxley TJ, Mocco J, Majidi S, Kellner CP, Shoirah H, Singh IP, et al. Large-Vessel Stroke as a Presenting Feature of Covid-19 in the Young. N Engl J Med. 2020 Apr 28;0(0):e60.

56. Avula A, Nalleballe K, Narula N, Sapozhnikov S, Dandu V, Toom S, et al. COVID-19 presenting as stroke. Brain Behav Immun [Internet]. 2020 Apr 28 [cited 2020 May 11]; Available from: https://www.ncbi.nlm.nih.gov/pmc/articles/PMC7187846/

57. González-Pinto T, Luna-Rodríguez A, Moreno-Estébanez A, Agirre-Beitia G, Rodríguez-Antiguedad A, Ruiz-Lopez M. Emergency Room Neurology in times of COVID- 19: Malignant Ischemic Stroke and SARS-COV2 Infection. Eur J Neurol [Internet]. [cited 2020 May 6];n/a(n/a). Available from: https://onlinelibrary.wiley.com/doi/abs/10.1111/ene.14286

58. Saiegh FA, Ghosh R, Leibold A, Avery MB, Schmidt RF, Theofanis T, et al. Status of SARS-CoV-2 in cerebrospinal fluid of patients with COVID-19 and stroke. J Neurol Neurosurg Psychiatry [Internet]. 2020 Apr 30 [cited 2020 May 10]; Available from: https://jnnp.bmj.com/content/early/2020/04/30/jnnp-2020-323522

59. Griffin DO, Jensen A, Khan M, Chin J, Chin K, Parnell R, et al. Arterial thromboembolic complications in COVID-19 in low risk patients despite prophylaxis. Br J Haematol [Internet]. [cited 2020 May 12];n/a(n/a). Available from: https://onlinelibrary.wiley.com/doi/abs/10.1111/bjh.16792

60. Association of British Neurologists. Association of British Neurologists Guidance on COVID-19 for people with neurological conditions, their doctors and carers [Internet]. [cited 2020 May 5]. Available from: https://www.neural.org.uk/wp-content/uploads/2020/03/25.3.20_ABN_Neurology_COVID-19_Guidance_v4.pdf

61. EANcore COVID-19 [Internet]. ean.org. [cited 2020 May 5]. Available from: https://www.ean.org/ean/eancore-covid-19

62. Temporary Emergency Guidance to US Stroke Centers During the COVID-19 Pandemic. Stroke. 2020 Apr;STROKEAHA.120.030023.

63. Qureshi AI, Abd-Allah F, Alsenani F, Aytac E, Borhani-Haghighi A, Ciccone A, et al. Management of acute ischemic stroke in patients with COVID-19 infection: Report of an international panel. Int J Stroke. 2020 May 3;17 47493020923234.

64. Müller-Barna P, Hubert GJ, Boy S, Bogdahn U, Wiedmann S, Heuschmann PU, et al. TeleStroke units serving as a model of care in rural areas: 10-year experience of the TeleMedical project for integrative stroke care. Stroke. 2014 Sep;45(9):2739–44.

65. Huang Y, Zhao N. Generalized anxiety disorder, depressive symptoms and sleep quality during COVID-19 epidemic in China: a web-based cross-sectional survey.:18.

66. Altena E, Baglioni C, Espie CA, Ellis J, Gavriloff D, Holzinger B, et al. Dealing with sleep problems during home confinement due to the COVID-19 outbreak: practical recommendations from a task force of the European CBT-I Academy. J Sleep Res [Internet]. 2020 Apr 4 [cited 2020 Apr 25]; Available from: http://doi.wiley.com/10.1111/jsr.13052

67. Mehta P, McAuley DF, Brown M, Sanchez E, Tattersall RS, Manson JJ. COVID-19: consider cytokine storm syndromes and immunosuppression. The Lancet. 2020 Mar;395(10229):1033–4.

68. HIV infection, hypercoagulability and ischaemic stroke in adults at the University Teaching Hospital in Zambia: a case control study. - PubMed - NCBI [Internet]. [cited 2020 May 10]. Available from: https://www.ncbi.nlm.nih.gov/pubmed/28521833

69. Nagel MA, Mahalingam R, Cohrs RJ, Gilden D. Virus Vasculopathy and Stroke: An Under-Recognized Cause and Treatment Target. Infect Disord Drug Targets. 2010 Apr 1;10(2):105–11.

70. Infectious causes of stroke | Request PDF [Internet]. ResearchGate. [cited 2020 May 10]. Available from: https://www.researchgate.net/publication/262784642_Infectious_causes_of_stroke

71. Baig AM, Khaleeq A, Ali U, Syeda H. Evidence of the COVID-19 Virus Targeting the CNS: Tissue Distribution, Host–Virus Interaction, and Proposed Neurotropic Mechanisms. ACS Chem Neurosci. 2020 Apr 1;11(7):995–8.

72. Finsterer J, Stollberger C. Causes of hypogeusia/hyposmia in SARS-CoV2 infected patients. J Med Virol [Internet]. [cited 2020 May 10];n/a(n/a). Available from: https://onlinelibrary.wiley.com/doi/abs/10.1002/jmv.25903

73. Paniz□Mondolfi A, Bryce C, Grimes Z, Gordon RE, Reidy J, Lednicky J, et al. Central Nervous System Involvement by Severe Acute Respiratory Syndrome Coronavirus □ 2 (SARS□CoV□2). J Med Virol. 2020 Apr 21;jmv.25915.

74. World Map - Simple [Internet]. MapChart. [cited 2020 May 13]. Available from: https://mapchart.net/world.html

